# Human genetics suggests differing causal pathways from *HMGCR* inhibition to coronary artery disease and type 2 diabetes

**DOI:** 10.1101/2025.02.10.25321990

**Authors:** Seongwon Hwang, Ville Karhunen, Ashish Patel, Sam Lockhart, Paul Carter, John Whittaker, Stephen Burgess

## Abstract

**Background:** Statins lower low-density lipoprotein cholesterol (LDL-C) and reduce the risk of coronary artery disease (CAD). However, they also increase the risk of type 2 diabetes (T2D).

**Methods:** We consider genetic variants in the region of the *HMGCR* gene, which encodes the target of statins, and their associations with downstream consequences of statins. We use various statistical methods to identify causal pathways influencing CAD and T2D, and investigate whether these are the same or different for the two diseases.

**Results:** Colocalization analyses indicated that LDL-C and body mass index (BMI) have distinct genetic predictors in this gene region, suggesting that they do not lie on the same causal pathway. Multivariable Mendelian randomization analyses restricted to variants in the *HMGCR* gene region revealed LDL-C and BMI as causal risk factors for CAD, and BMI as a causal risk factor for T2D, but not LDL-C. A Bayesian model averaging method prioritized BMI as the most likely causal risk factor for T2D, and LDL-C as the second most likely causal risk factor for CAD (behind ubiquinone). Colocalization analyses provided consistent evidence of LDL-C colocalizing with CAD, and BMI colocalizing with T2D; evidence was inconsistent for colocalization of LDL-C with T2D, and BMI with CAD.

**Conclusions:** Our analyses suggest cardiovascular and metabolic consequences of statin usage are on different causal pathways, and hence could be influenced separately by targeted interventions. More broadly, our analysis workflow offers potential insights to identify pathway-specific causal risk factors that could provide possible repositioning or refinement opportunities for existing drug targets.

**Key messages:**

- We performed colocalization and cis-multivariable Mendelian randomization using genetic association data for variants in the *HMGCR* gene region to investigate causal pathways influencing coronary artery disease (CAD) and type 2 diabetes (T2D)
- Our analyses suggest that the impact of HMGCR inhibition on CAD risk is mediated by both low-density lipoprotein cholesterol (LDL-C) and body mass index (BMI), whereas for T2D, risk was mediated via BMI but not LDL-C.
- Our results suggest the possibility that targeted treatments could be developed to inhibit HMGCR in a more specific way that lowers CAD risk without increasing T2D risk.

## INTRODUCTION

Statins are a well-known class of medications that inhibit 3-hydroxy-3-methyl-glutaryl-coenzyme A reductase (HMGCR), the rate-controlling enzyme in the mevalonate pathway.^1^ This is a metabolic pathway that synthesizes cholesterol and other organic chemicals. Statins are well-established by randomized clinical trials to lower low-density lipoprotein cholesterol (LDL-C) and reduce the risk of coronary artery disease (CAD) making them a cornerstone of cardiovascular disease preventive therapy.^2^ Similar reductions in CAD risk have been observed for other LDL-C lowering agents.^3–5^ However, statins have also been shown in trials^6^ and population-based studies^7,8^ to increase incident risk of type 2 diabetes (T2D), and in some cases to worsen glycaemic control^9^ and progression to insulin requirement^10^ in established diabetics. Despite this, statins provide an overall benefit with respect to microvascular and macrovascular cardiovascular disease,^11^ although a therapeutic option which isolates the LDL-C lowering effect of statins from its diabetogenic effects would be clinically desirable.

Genetic variants can be used to predict the results of clinical trials using a technique known as Mendelian randomization.^12^ Individuals with certain genetic variants in the *HMGCR* gene region have a natural predisposition to increased inhibition of the mevalonate pathway that is analogous to taking a low-dose statin.^13^ As genetic variants are inherited at random conditional on the parental genotype according to Mendel’s laws, epidemiological associations of variants in the *HMGCR* gene should reflect the downstream consequences of taking statins.^14^ Empirical investigations have suggested that genetic associations in a well-mixed and homogeneous population are not systemically affected by confounding, and so the associations should be a reliable guide as to the effects of statins.^15,16^ Indeed, variants in the *HMGCR* gene region that associate with higher LDL-C are also associated with greater risk of CAD^17^ and lower risk of T2D^18^, in line with clinical trials.

However, clinical trials of other LDL-C lowering agents (including PCSK9 inhibitors^19^, NPC1L1 inhibitors^20^, and bempedoic acid^21^) have not demonstrated increases in T2D risk, and genetic associations of variants in corresponding gene regions show between-region heterogeneity in their associations with T2D risk.^22,23^ In contrast, genetic associations with CAD risk are proportional to the genetic associations with LDL-C for variants in different drug-mimicking gene regions.^24,25^ This suggests that LDL-C may not be driving the increases in T2D risk observed in statin trials.

The aim of this investigation is to explore the mechanisms linking the target of statins to CAD and T2D using multiple genetic variants in the *HMGCR* gene region. We use various statistical techniques to investigate causal pathways, including colocalization, multivariable Mendelian randomization, and Bayesian model averaging. We discuss the implications of these analyses for lipid lowering treatment strategies in practice.

## METHODS

### Study overview

We consider associations of genetic variants in the *HMGCR* gene region with risk factors that have been demonstrated to be consequences of statin usage, and with CAD and T2D risk. For each pair of risk factors, we perform colocalization to investigate whether the risk factor association signals are driven by the same or different variants. For pairs of risk factors driven by different variants, we conduct multivariable Mendelian randomization analyses with CAD and T2D as outcomes. We also use a Bayesian approach to calculate the posterior probabilities of all possible risk factor models, and hence identify the most statistically plausible risk factors affecting CAD and T2D. While we would be cautious about an overly literalistic interpretation of the statistical results that we present here, strong differences in results for CAD versus T2D as the outcome would be indicative of different mechanisms by which *HMGCR*-regulated interventions on the mevalonate pathway may affect these two diseases. An overview of analyses is displayed as **Figure 1**.

**Figure 1.**
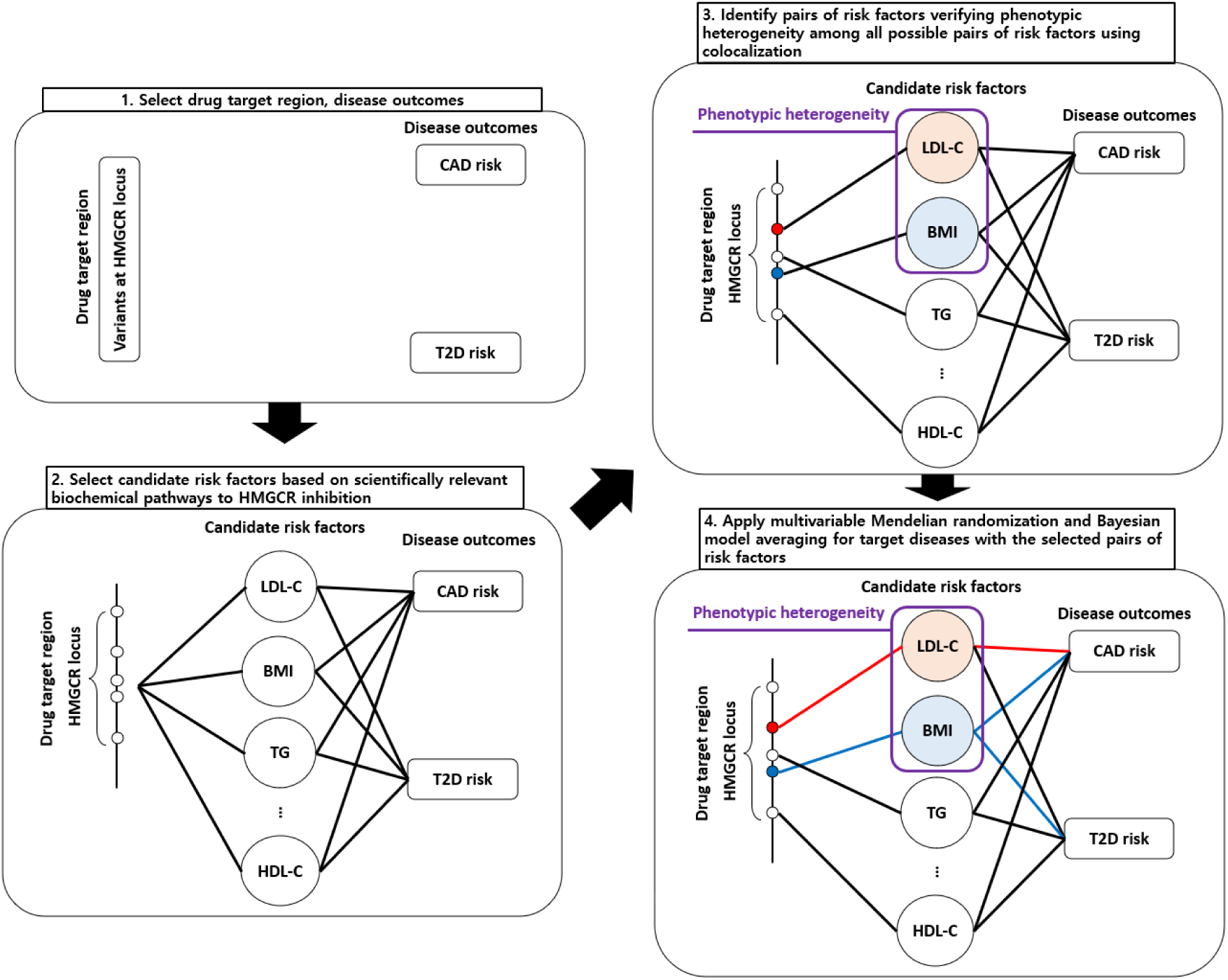
Overall flowchart of our workflow. Abbreviations: CAD: coronary artery disease, LDL-C: low-density lipoprotein cholesterol, HDL-C: high density lipoprotein cholesterol, BMI: body mass index, TG: triglyceride, T2D: type 2 diabetes, *HMGCR*: 3-hydroxy-3-methyl-glutaryl-coenzyme A reductase.

### Risk factors

We selected 16 candidate risk factors based on scientifically relevant biochemical pathways to *HMGCR* inhibition and availability of summarized genetic association data. These were: LDL-C, high-density lipoprotein cholesterol (HDL-C), triglyceride, sterol, ubiquinone, cortisol, testosterone, estradiol, aldosterone, 25-hydroxyvitamin D, bile acids, body mass index (BMI), leptin, acute insulin response, fasting insulin, and fasting glucose. This choice of variables provides comprehensive coverage of major lipids, steroid hormones (which are synthesized from cholesterol), and metabolic traits, as well as other cholesterol-derived compounds (25-hydroxyvitamin D and bile acids). Datasets used to obtain estimates of genetic associations with these risk factors are listed in **Supplementary Table 1**.

### Outcomes

Genetic associations with CAD were obtained from a large genome-wide association study comprising 181,522 cases and 984,168 controls predominantly of European ancestry.^26^ Genetic associations with T2D were obtained from the Diabetes Meta-Analysis of Trans-Ethnic association studies (DIAMANTE) Consortium analysis of 80,154 cases and 853,816 controls of European ancestry.^27^

### Colocalization

Colocalization is a statistical method to distinguish between two scenarios at a given gene region for a pair of traits: 1) the traits have shared genetic predictors (known as colocalization, and consistent with the traits being on the same causal pathway), and 2) the traits have distinct genetic predictors (known as non-colocalization, and consistent with the traits being on different causal pathways).^28^ While in typical applications of Mendelian randomization, we want to see colocalization between the exposure and outcome, in this case we are interested in finding pairs of traits that have distinct genetic predictors because this enables multivariable Mendelian randomization analyses using the two traits as exposures; if two risk factors have distinct (i.e. not proportional) genetic predictors, a multivariable model can distinguish between their effects.

Colocalization analyses were performed using four methods, which address slightly different questions and/or make slightly different assumptions. The coloc method considers evidence for five hypotheses.^29^ The critical hypotheses for this work are H3 (distinct causal variants for each trait; that is, non-colocalization) and H4 (shared causal variant for both traits; that is, colocalization). Other hypotheses are that there is only a causal variant for trait 1 (H1), only for trait 2 (H2), or no causal variants for either of the traits (H0). The coloc-SuSiE (sum of single effects) method is an extension of the coloc method that allows each trait to have multiple causal variants, represented by distinct credible sets.^30^ It considers whether or not there is colocalization for each pair of detected causal variants using the same set of hypotheses as coloc.

The proportional colocalization method assesses whether genetic associations with one trait are proportional with those of the other trait or not.^31^ Proportionality suggests that the two traits are on the same causal pathway. The prop.coloc method reports two p-values: the Lagrange multiplier (LM) test, which tests the null hypothesis that the proportionality constant is zero, and the proportionality test, which tests the null hypothesis that the genetic associations are proportional. Non-colocalization is concluded when the LM test and proportionality tests both reject the null hypothesis. If the LM test rejects the null but the proportionality test does not, then there is no evidence to reject the colocalization hypothesis.

The colocPropTest method also assesses proportionality in genetic associations.^32^ It does this by taking pairs of variants in turn, assessing proportionality in genetic associations using a heterogeneity test for that pair, and using false discovery rates to account for multiple testing. If there is sufficient evidence, the null hypothesis of colocalization is rejected in favour of the alternative hypothesis of non-colocalization.

Colocalization analyses were performed for two purposes: first, we investigate colocalization between exposures to find pairs of exposures with separate genetic predictors, and second, we investigate colocalization between exposures used in our multivariable Mendelian randomization analyses with disease outcomes, to validate their status as causal risk factors.

### Multivariable Mendelian randomization

Standard Mendelian randomization takes genetic predictors of a single exposure, and assesses whether genetic predictors of that exposure (or equivalently, genetically-predicted levels of the exposure) are associated with the outcome in a univariable regression model. Under the instrumental variable assumptions^12^, an association between genetically-predicted levels of the exposure and the outcome is indicative of a causal effect of the exposure on the outcome.

Multivariable Mendelian randomization considers multiple related exposures, and assesses whether genetically-predicted levels of each exposure are conditionally associated with the outcome in a multivariable regression model.^33^ The motivation is that it can be difficult to find genetic variants that associate uniquely with a single exposure. Multivariable Mendelian randomization allows genetic variants to associate with any or all of the exposures in the model. Under the multivariable instrumental variable assumptions, a conditional association between genetically-predicted levels of an exposure and the outcome is indicative of a direct causal effect of that exposure on the outcome.^34^

In our example, genetic variants in the *HMGCR* gene region are associated with multiple risk factors that can be used as separate exposures in multivariable Mendelian randomization provided that their genetic associations are not proportional (as this would lead to collinearity in the regression model). This method can identify the proximal causal risk factors for an outcome. We perform multivariable Mendelian randomization for pairs of risk factors that have strong evidence for having non-proportional genetic associations (i.e. evidence for non-colocalization in the framework above).^35^ We refer to risk factors with non-proportional genetic associations as displaying “phenotypic heterogeneity”.

### Bayesian model averaging

Rather than considering pairs of risk factors separately, the Mendelian randomization Bayesian model averaging (MR-BMA)^36^ method allows the consideration of all risk factors in a model averaging framework. We include all risk factors displaying some evidence of phenotypic heterogeneity in this analysis. We perform Mendelian randomization for each possible model (that is, each subset of risk factors): each risk factor alone, each pair of risk factors, each triple of risk factors, and so on. For a small number of risk factors, this process can consider all models; for a larger number of risk factors, a stochastic search strategy is deployed. Each set of risk factors is assigned a Bayes factor depending on how well the data fit the model, and these are used to calculate the posterior probability of each model (each subset of risk factors). We also calculate the marginal inclusion probability (MIP) for each risk factor, representing the sum of posterior probabilities for models containing that risk factor. This provides a ranking of risk factors based on their likelihood of being a causal risk factor for the outcome.

### Statistical analyses

All analyses used genetic variants located within 10k base pairs of the *HMGCR* coding region (chromosome 5, positions 74,632,154-74,659,826 on build hg19). We consider a narrow range of variants around the coding region to ensure as far as possible that any genetic associations relate to inhibition of *HMGCR* and not to unrelated pathways.

Colocalization analyses were performed using coloc^37^ and coloc-SuSiE^30^ using default colocalization priors, prop-coloc-cond^31^ with a pruning level of 0.4, and colocPropTest^38^, and otherwise with default settings. When coloc-SuSiE finds multiple causal variants, we display results for the lead pair of credible sets in our plots, and provide full results in a separate table.

Multivariable Mendelian randomization analyses were implemented using the multivariable principal component generalized method of moments (MV-PC-GMM), which performs dimension reduction to reduce a large number of correlated variants to a manageable number of orthogonal principal components. This method was chosen as pruning strategies for *cis*-Mendelian randomization can be inefficient, particularly for multivariable Mendelian randomization as we require conditionally strong genetic predictors of each risk factor. The method is implemented in the mr_mvpcgmm function in the MendelianRandomization package with the random effects option. We used a threshold of 99% of variance in the matrix of genetic associations with the exposure for selecting principal components, allowing for random-effects heterogeneity, and using variant correlation estimates obtained from 367,703 European ancestry UK Biobank participants. To assess the sensitivity of the results to the proportion of variance explained, we perform a sensitivity analysis to compare results with different numbers of principal components.

MR-BMA was performed using default settings, including an independent prior inclusion probability of 0.1 for each risk factor.

Where required, colocalization analyses used linkage disequilibrium matrices estimates in European ancestry UK Biobank participants.

All analyses were performed in the R software environment (version 4.0.2), using packages susieR (version 0.12.35), coloc (version 5.2.3), colocPropTest (version 0.9.1), prop.coloc (version 1.1.0), MendelianRandomization (version 0.9.0), and mrbma (version 0.1.0).

## RESULTS

### Colocalization to identify phenotypic heterogeneity

Results from analyses of pairs of risk factors for the proportional colocalization and coloc-SuSiE/coloc methods are displayed in **Figure 2**. We show results for each risk factor that rejected at least one proportional colocalization test. We used the coloc-SuSiE method preferentially, and coloc in cases where coloc-SuSiE did not find a credible set of causal variants. Evidence for phenotypic heterogeneity was inferred from rejection (p<0.05) of the proportionality and LM tests in the proportional colocalization method, and a high value (>50%) for the non-colocalization hypothesis (H3) in coloc-SuSiE or coloc. This was only observed for one pair of traits: LDL-C and BMI. Results from all credible sets using coloc-SuSiE between LDL-C and BMI are reported in **Supplementary Table 2**; for each pair of credible sets, the posterior probability of H3 (PP-H3) was >0.84.

**Figure 2.**
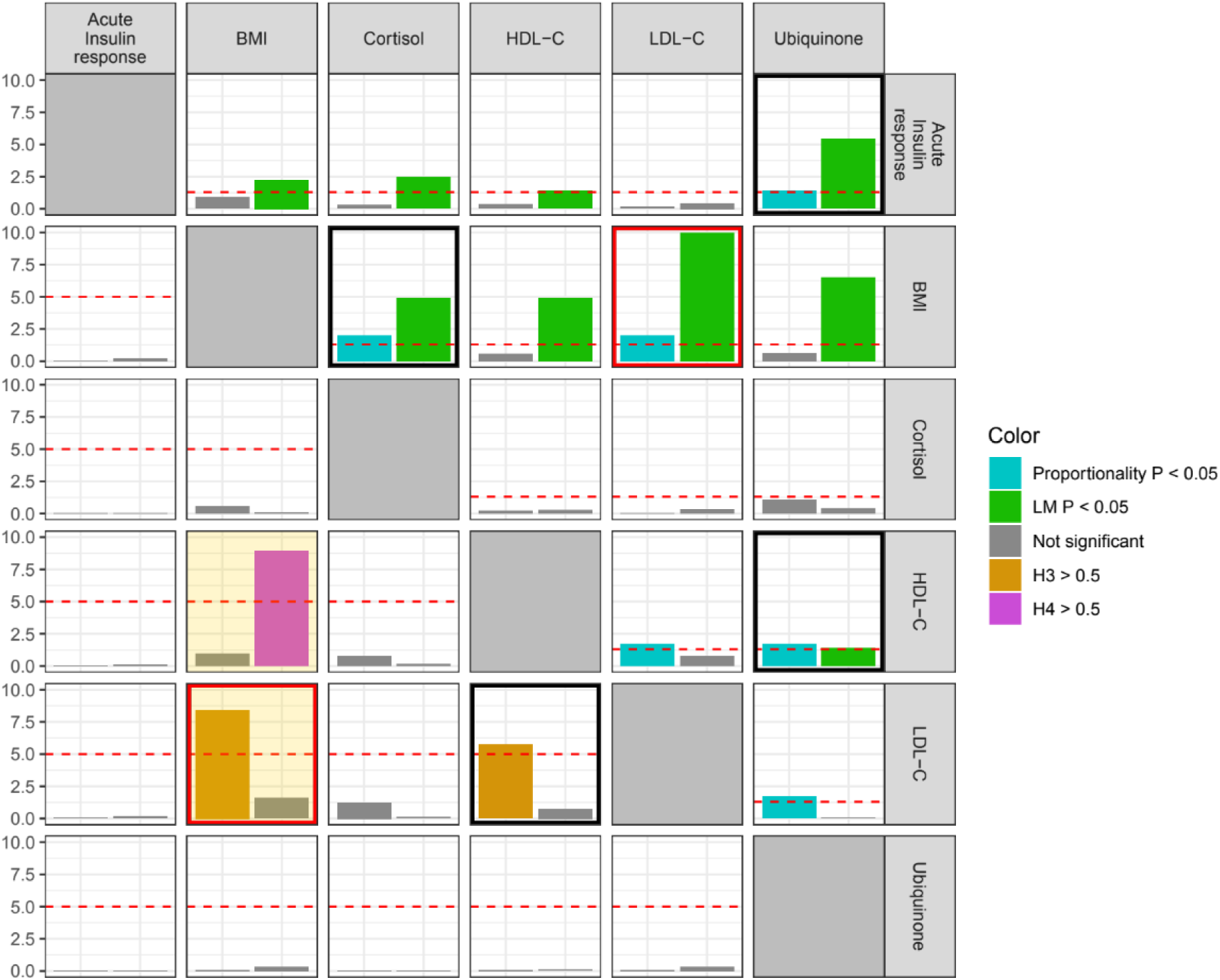
Heatmap of colocalization results from proportional colocalization and coloc-SuSiE/coloc methods. Top-right quadrant displays results from proportional colocalization method (prop-coloc-cond). Bars represent negative log_10_-transformed p-values for the proportionality test (cyan) and the Lagrange multiplier (LM) test (green). Phenotypic heterogeneity is indicated when both tests reject the null hypothesis (p<0.05, equivalent to - log_10_p>1.3, red horizontal line). Bottom-left quadrant displays results from coloc-SuSiE (orange-shaded) or coloc methods. Coloc-SuSiE was used preferentially, coloc was used in cases where coloc-SuSiE did not find a credible set of causal variants. Bars represent posterior probability percentage divided by 10 (that is, 10.0 represents 100%) for non-colocalization (H3, orange) or colocalization (H4, magenta). Phenotypic heterogeneity is indicated when the posterior probability for H3 is above 50% (red horizontal line). A black box indicates evidence for phenotypic heterogeneity from each method, a red box indicates evidence from both frameworks. Only traits with evidence of rejecting at least one colocalization test are displayed.

To validate this further, we conducted the colocPropTest method, which also indicated non-proportionality in the genetic associations with BMI and LDL-C (**Supplementary Figure 1**). We therefore focus on this pair in multivariable Mendelian randomization analyses. We note that this pair comprises one lipid trait and one metabolic trait.

### Multivariable Mendelian randomization for the *HMGCR* gene region

*cis*-Multivariable Mendelian randomization results are displayed in **Figure 3**. Mutually adjusted genetically-predicted levels of LDL-C and BMI were each independently associated with CAD risk: odds ratio (OR) 2.26 (95% confidence interval [CI]: 1.46, 3.50; p=0.0003) per 1 standard deviation increase in LDL-C, and OR 4.02 (95% CI: 1.09, 14.78; p=0.04) per 1 kg/m^2^ increase in BMI. In contrast, genetically-predicted levels of BMI were conditionally associated with T2D risk: OR 6.60 (95% CI: 1.28, 34.16; p=0.03), but there was no clear association for genetically-predicted levels of LDL-C: OR 1.17 (95% CI: 0.67, 2.03; p=0.58). Analyses were based on 119 variants for CAD and 96 variants for T2D; in both cases, the top 3 principal components explained 99% variance in the gene-exposure association matrix. Conditional F statistics were 10.6 for LDL-C and 10.6 for BMI on CAD, and 11.1 for LDL-C and 11.0 for BMI on T2D. Similar results were obtained in sensitivity analyses using different numbers of principal components (**Supplementary Table 3**).

**Figure 3.**
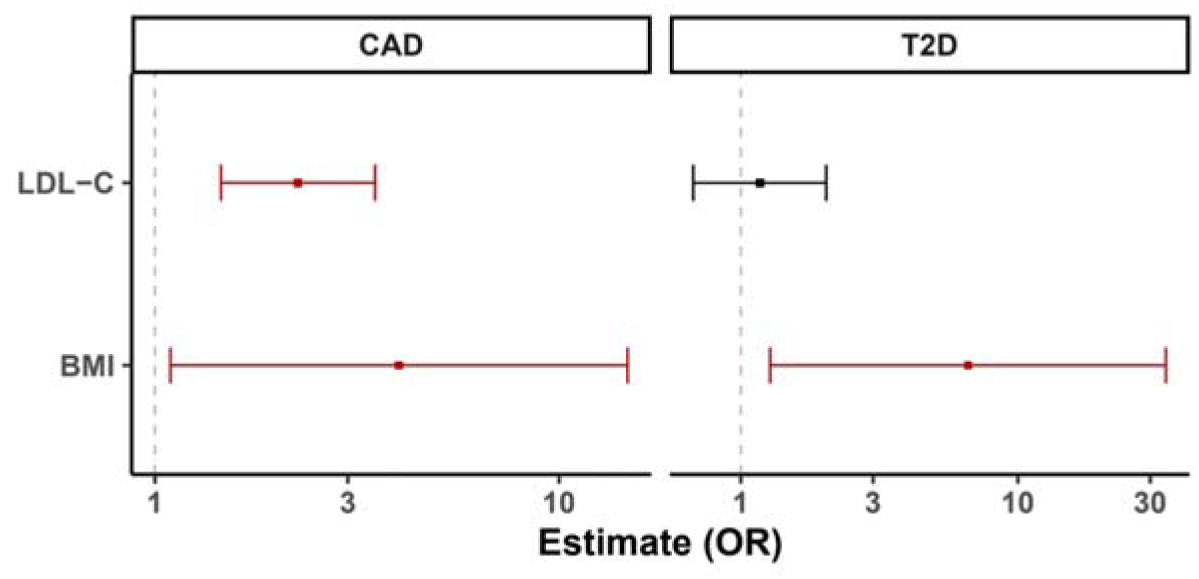
Multivariable Mendelian randomization results. Estimates represent mutually adjusted odds ratios for coronary artery disease (CAD, left) and Type 2 diabetes (T2D, right) per 1 standard deviation increase in genetically-predicted low-density lipoprotein cholesterol (LDL-C) or per 1 kg/m^2^ increase in body mass index (BMI) from multivariable analyses restricted to genetic variants in the *HMGCR* locus.

### Colocalization to validate Mendelian randomization findings

We performed colocalization analyses to support or refute our Mendelian randomization findings. If an exposure colocalizes with an outcome, this increases our confidence that the exposure and outcome are on the same causal pathway. Results are summarized in **Table 1**, and full results from coloc-SuSiE when there were multiple credible sets are provided in **Supplementary Table 4**.

**Table 1.**
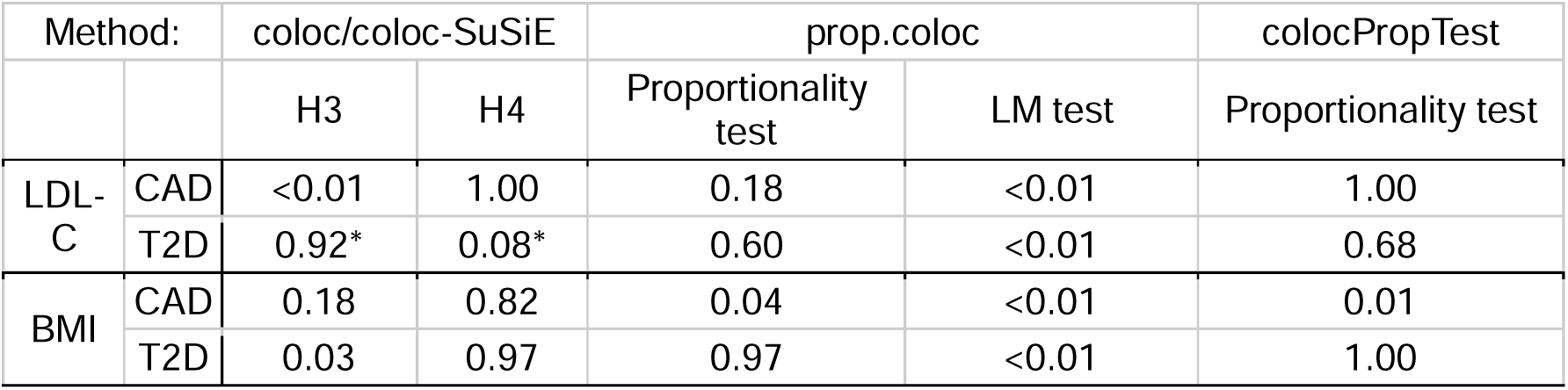
Summary of results from colocalization analyses to validate Mendelian randomization.

For LDL-C and CAD, there was strong evidence for colocalization from the coloc method (posterior probability of H4 [PP-H4] = 1.00), and no evidence to reject colocalization in either of the proportional colocalization methods. For LDL-C and T2D, there was strong evidence for non-colocalization from the coloc-SuSiE method, with PP-H3 > 0.92 for all four pairs of credible sets. However, the proportional colocalization methods did not reject colocalization.

For BMI and CAD, there was mixed evidence, with coloc-SuSiE favouring colocalization (PP- H3 = 0.18, PP-H4 = 0.82), but the proportional colocalization methods both rejecting colocalization. For BMI and T2D, there was strong evidence for colocalization from the coloc method (PP-H4 = 0.97), and no evidence to reject colocalization in either of the proportional colocalization methods.

In summary, we found consistent evidence in the *HMGCR* gene region to validate LDL-C as a causal risk factor for CAD, and BMI as a causal risk factor for T2D, and inconsistent evidence for BMI as a causal risk factor for CAD, and LDL-C as a causal risk factor for T2D. Together, our suggest that BMI raising variants in the *HMGCR* region increase T2D risk, whereas LDL-C lowering variants reduce CAD but do not affect T2D risk. This is consistent with the effects of statins on CAD and T2D being mediated by distinct molecular mechanisms.

### Bayesian model averaging

Results from the MR-BMA method are shown in **Table 2**. For CAD, the top ranking model was the model containing ubiquinone as the single risk factor, and the top ranking risk factor by posterior inclusion probability was ubiquinone (MIP = 27.68%) followed by LDL-C (MIP = 25.04%). We note that reduced ubiquinone is a direct molecular consequence of HMGCR inhibition, and hence this is likely not a competing risk factor to LDL-C, but an upstream trait on shared causal pathways. For T2D, the top ranked model was BMI as a single risk factor, and BMI was the top ranked risk factor (MIP = 54.64%).

**Table 2.** Top ranking models and risk factors from Mendelian randomization Bayesian model averaging (MR-BMA) method: Risk factors showing phenotypic heterogeneity in **Figure 2** are used as inputs in this method.

| Coronary artery disease |  |  |
| --- | --- | --- |
| Rank | Top models (subsets of risk factors) | Posterior probability (%) |
| 1 | Ubiquinone | 19.11% |
| 2 | Acute Insulin response | 15.52% |
| 3 | Cortisol | 14.24% |
| 4 | LDL-C | 9.07% |
| 5 | HDL-C | 7.42% |
| Rank | Top risk factors | Marginal inclusion probability (%) |
| 1 | Ubiquinone | 27.68% |
| 2 | LDL-C | 25.04% |
| 3 | Acute Insulin response | 23.61% |
| 4 | Cortisol | 23.17% |
| 5 | BMI | 17.47% |
| Type 2 diabetes |  |  |
| Rank | Top models (subsets of risk factors) | Posterior probability (%) |
| 1 | BMI | 34.65% |
| 2 | HDL-C | 19.94% |
| 3 | Ubiquinone | 5.05% |
| 4 | Acute Insulin response | 4.82% |
| 5 | BMI, HDL-C | 4.81% |
| Rank | Top risk factors | Marginal inclusion probability (%) |
| 1 | BMI | 54.64% |
| 2 | HDL-C | 34.55% |
| 3 | Ubiquinone | 14.60% |
| 4 | Acute Insulin response | 14.46% |
| 5 | Cortisol | 13.55% |

## DISCUSSION

In this investigation, we performed various statistical analyses using genetic association data for variants in the *HMGCR* gene region to investigate causal pathways influencing CAD and T2D. Colocalization analyses indicated that there are distinct genetic predictors of LDL-C and BMI, a finding suggesting that these traits are influenced by separate causal pathways. A targeted multivariable Mendelian randomization analyses including these two traits gave differing results for the two diseases: for CAD, it suggested that LDL-C and BMI were causal mediators of disease risk, and for T2D, it suggested that BMI was a mediator and LDL-C was not. A multivariable analysis including a wider range of risk factors gave similar results: it prioritized LDL-C as the second most likely causal risk factor for CAD (behind ubiquinone, a biomarker of HMGCR inhibition), whereas it prioritized BMI as the most likely causal risk factor for T2D. Colocalization analyses indicated consistent evidence for LDL-C as colocalizing with CAD, and BMI colocalizing with T2D. Although we would be cautious about an overly literal interpretation of these results, it is clear that the results are very different for the two diseases. Together our results suggest that HMGCR inhibition by statins may affect distinct causal pathways to cause T2D and protect against CAD.

Our findings are consistent with previous results from both experimental and observational studies. Randomized clinical trials have shown that LDL-C lowering using statins reduces CAD risk ^39^. Moreover, multiple *in vivo* and clinical studies have substantiated a causal relationship between obesity and T2D.^40^ Randomized trials have also shown that the weight loss drug semaglutide lowers risk of cardiovascular disease.^41^ Aside from clinical evidence, several Mendelian randomization analyses have been performed to verify the causal link between LDL-C and risk for CAD^42^, the causal link between BMI and T2D^43^, and the causal link between BMI and CAD^44^, including analyses based on variants in the *HMGCR* locus.^22^ However, previous analyses have not investigated the causal pathways underlying these signals, particularly using multivariable methods that are able to distinguish between pathways.

While statins have clear benefits in terms of reducing CAD risk, they have unwanted effects in terms of increasing T2D risk. Our results suggest that these effects are on different causal pathways, raising the possibility that targeted treatments could be developed to inhibit *HMGCR* in a more specific way that lowers CAD risk without increasing T2D risk. We attempted to provide greater insight into the mechanisms of action using data on *HMGCR* gene expression, but we did not find strong evidence for colocalization between gene expression and known downstream consequences of statins, such as LDL-C and BMI. Other investigators considering genetic proxies of drug targets have also found that gene expression is not always a reliable guide of the mechanism of action^45^, and a recent review recommended using downstream traits to calibrate and guide choice of genetic variants where possible, rather than gene expression or protein levels.^46^ However, while our analyses are not able to pinpoint how this could be achieved, it should encourage drug manufacturers to consider different mechanisms of action and modalities of treatment in drugs that target the mevalonate pathway, and that deeper functional characterization of these genetic effects is warranted. It also suggests that increases in T2D risk may not be seen equally for all LDL-C lowering targets. More generally, our investigation provides an example of how drug target Mendelian randomization can provide translational insights into drug development.

Our investigation has limitations. As with all Mendelian randomization analyses, we rely on the validity of the genetic variants as instrumental variables. More specifically, we rely on the genetic variants as satisfying the gene—environment equivalence assumption for statins^47^; that is, the genetic variants influence traits and outcomes in a similar way to statins. This would not be satisfied if genetic variants have pleiotropic effects on other traits, or associations with other traits arising from linkage disequilibrium with a variant in another gene region, or from population stratification. We have minimized the possibility of this by restricting our analyses to variants in a narrow region around the *HMGCR* gene; however, we cannot fully discount this possibility. Our estimates may suffer from weak instrument bias. Weak instrument bias is particularly pervasive in multivariable Mendelian randomization, as we require not only strong genetic predictors of all the traits in the model, but some degree of independence in these genetic predictors.^48^ While the generalized method of moments is less sensitive to weak instruments than some other methods^49^, some bias may remain. Although genetic variants mimic interventions on drug targets in many aspects, genetic associations represent life-long, small changes differences in risk factor levels, whereas pharmacological interventions are typically shorter in duration, but greater in magnitude.^50^ Finally, our analyses were performed in predominantly European ancestry populations. While we would not expect strong differences in biological effects between ancestry groups, there are differences between populations in terms of disease prevalence, risk factor distributions, and response to metabolic changes^51^ that could lead to different findings for different ancestry groups.

In conclusion, we have found evidence from human genetics that different consequences of statin usage are on different causal pathways, and hence could be influenced separately by targeted interventions. Future investigations into drugs that inhibit the *HMGCR* pathway should investigate whether lipid lowering can be achieved without metabolic dysfunction.

## Data Availability

Datasets used to obtain estimates of genetic associations with these risk factors are listed in Supplementary Table 1.

https://sw4rsw4r.github.io/HMGCR-Causality/HMGCR-Causality.html

## Acknowledgements

This research has been conducted using the UK Biobank Resource under Application Number 98032.

## Code availability

R scripts to apply all methods used in this manuscript including multivariable Mendelian randomization, colocalization, and Bayesian model averaging are available at https://github.com/sw4rsw4r/HMGCR-Causality and R markdown to allow the reader to reproduce all results, figures and tables is available at https://sw4rsw4r.github.io/HMGCR-Causality/HMGCR-Causality.html.

## Ethics declarations

Our study only uses publicly available summarized data, and so does not require specific ethical approval. Ethical approval for the original studies can be found in the relevant references.

---

**Supplementary Figure 1.**
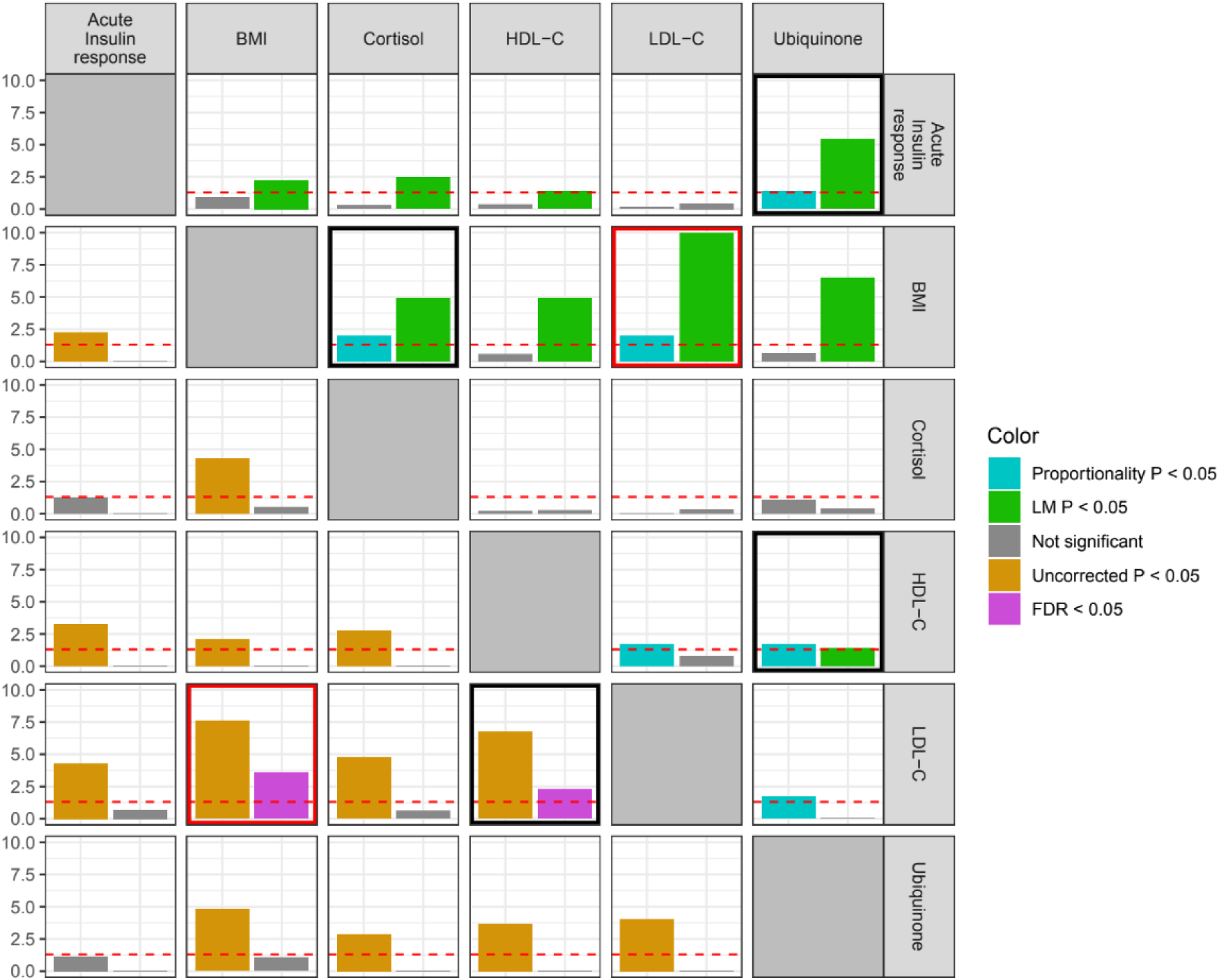
Heatmap of colocalization results from proportional colocalization and colocPropTest methods. Top-right quadrant displays results from proportional colocalization method (prop-coloc-cond). Bars represent negative log_10_- transformed p-values for the proportionality test (cyan) and the Lagrange multiplier (LM) test (green). Phenotypic heterogeneity is indicated when both tests reject the null hypothesis (p<0.05, equivalent to -log_10_p>1.3, red horizontal line). Bottom-left quadrant displays results from colocPropTest method. Bars represent negative log_10_-transformed p-values for the proportionality test: uncorrected p-value (orange), and false-discovery rate (FDR) corrected p-value (magenta). Phenotypic heterogeneity is indicated when the FDR corrected p-value is less than 0.05 (horizontal red line). A black box indicates evidence for phenotypic heterogeneity from each method, a red box indicates evidence from both frameworks. Only traits with evidence of rejecting at least one colocalization test are displayed.

**Supplementary Table 1.** List of risk factors and outcomes.

| Phenotype | Study reference | GWAS Catalog dataset ID or weblink |
| --- | --- | --- |
| Acute insulin response | Wood et al, Diabetes 2017; 66:2296-2309 | GCST004575 |
| Fasting insulin | Manning et al, Nat Genet 2012; 44:659-669 | GCST005185 |
| Fasting glucose | Manning et al, Nat Genet 2012; 44:659-669 | GCST005186 |
| BMI | Pulit et al, Hum Mol Genet. 2019; 28:166-174 | GCST009004 |
| LDL-C | Graham et al, Nature 2021; 600:675-679 | GCST90239658 |
| HDL-C | Graham et al, Nature 2021; 600:675-679 | GCST90239652 |
| Triglycerides | Graham et al, Nature 2021; 600:675-679 | GCST90239664 |
| Leptin | Yaghootkar et al, Diabetes. 2020; 69:2806-2818 | GCST90007310 |
| Sterol | Harshfield et al, BMC Medicine 2021; 19:232 | GCST90060133 |
| Cortisol | Chen et al, Nat Genet 2023; 55:44-53 | GCST90200378 |
| Testosterone | Leinonen et al, Communications Medicine. 2023; 3:4 | GCST90239820 |
| Estradiol | Schmitz et al, J Clin Endocrinol Metab. 2021;106: e4471-e4486 | GCST90020092 |
| Vitamin D | Revez et al, Nat Commun 2020; 11:1647 | GCST90000618 |
| Bile acid | Harshfield et al, BMC Medicine 2021; 19:232 | GCST90060135 |
| Aldosterone | Dennis et al, Genome Med 2021; 13:6 | GCST90012609 |
| Ubiquinone | Cadby et al, Nat Commun 2022; 13:3124 | GCST90024608 |
| CAD | Aragam et al, Nat Genet. 2022;54: 1803–1815 | GCST90132314 |
| T2D | Mahajan et al, Nat Genet. 2022;54: 560–572 | https://www.diagram-consortium.org/downloads.html |

**Supplementary Table 2.** Full coloc-SuSiE results for LDL-C and BMI: numbers in bold are used for plotting in **Figure 2**.

| LDL-C and BMI |  |  | Posterior probability for hypothesis: |  |  |  |  |
| --- | --- | --- | --- | --- | --- | --- | --- |
| Number of SNPs | Lead SNP for LDL-C | Lead SNP for BMI | H0 | H1 | H2 | H3 | H4 |
| **148** | **rs12916** | **rs3843480** | **<0.01** | **<0.01** | **<0.01** | **0.84** | **0.16** |
| 148 | rs72633963 | rs3843480 | <0.01 | <0.01 | <0.01 | 1.00 | <0.01 |
| 148 | rs184049365 | rs3843480 | <0.01 | <0.01 | <0.01 | 1.00 | <0.01 |
| 148 | rs7717396 | rs3843480 | <0.01 | <0.01 | <0.01 | 1.00 | <0.01 |
| 148 | rs113586903 | rs3843480 | <0.01 | <0.01 | <0.01 | 1.00 | <0.01 |

There are five credible sets in coloc-SuSiE for LDL-C and one for BMI. Colocalization between each pair of sets indicated evidence for distinct causal variants (H3).

**Supplementary Table 3.** Sensitivity analysis for multivariable Mendelian randomization investigation varying the number of principal components.

| Number of principal components | P-value (CAD) |  | Number of principal components | P-value (T2D) |  |
| --- | --- | --- | --- | --- | --- |
|  | BMI | LDL-C |  | BMI | LDL-C |
| 3 (99%) | 0.036 | <0.001 | 3 (99%) | 0.024 | 0.581 |
| 4 | 0.039 | <0.001 | 4 | 0.027 | 0.708 |
| 5 (99.9%) | 0.029 | <0.001 | 5 (99.9%) | 0.017 | 0.615 |
| 6 | 0.01 | <0.001 | 6 | 0.002 | 0.184 |
| 7 | 0.009 | <0.001 | 7 | 0.002 | 0.197 |
| 13 (99.99%) | 0.004 | <0.001 | 12 (99.99%) | 0.001 | 0.780 |

For each number of principal components, multivariable Mendelian randomization indicated a causal role of LDL-C and BMI for coronary artery disease (CAD), and a causal role of BMI (but not LDL-C) for type 2 diabetes (T2D).

**Supplementary Table 4.** Full coloc-SuSiE results for LDL-C and T2D.

| LDL-C and T2D |  |  | Posterior probability for hypothesis: |  |  |  |  |
| --- | --- | --- | --- | --- | --- | --- | --- |
| Number of SNPs | Lead SNP for LDL-C | Lead SNP for T2D | H0 | H1 | H2 | H3 | H4 |
| 111 | rs12916 | rs7733436 | <0.01 | <0.01 | <0.01 | 0.92 | 0.08 |
| 111 | rs138091235 | rs7733436 | <0.01 | <0.01 | <0.01 | 1.00 | <0.01 |
| 111 | rs4703671 | rs7733436 | <0.01 | <0.01 | <0.01 | 0.98 | 0.01 |
| 111 | rs78585310 | rs7733436 | <0.01 | <0.01 | <0.01 | 0.99 | <0.01 |

There are four credible sets in coloc-SuSiE for LDL-C and one for T2D. Colocalization between each pair of sets indicated evidence for distinct causal variants (H3).

